# Changes of evening exposure to electronic devices during the COVID-19 lockdown affect the time course of sleep disturbances

**DOI:** 10.1101/2020.10.20.20215756

**Authors:** Federico Salfi, Giulia Amicucci, Domenico Corigliano, Aurora D’Atri, Lorenzo Viselli, Daniela Tempesta, Michele Ferrara

## Abstract

**Study Objectives:** During the COVID-19 lockdown, there was a worldwide increase in electronic devices’ daily usage. The exposure to backlit screens before falling asleep leads to negative consequences on sleep health through its influence on the circadian system. We investigated the relationship between the changes in evening screen exposure and the time course of sleep disturbances during the home confinement period due to COVID-19.

**Methods:** 2123 Italians were longitudinally tested during the third and the seventh week of lockdown. The web-based survey evaluated sleep quality and insomnia symptoms through the Pittsburgh Sleep Quality Index and the Insomnia Severity Index. During the second assessment, respondents reported the changes in the backlit screen exposure in the two hours before falling asleep.

**Results:** Participants who increased electronic device usage showed decreased sleep quality, exacerbated insomnia symptoms, reduced sleep duration, higher sleep onset latency, and delayed bedtime and rising time. In this subgroup, the prevalence of poor sleepers and clinical insomniacs increased. Conversely, respondents reporting decreased screen exposure exhibited improved sleep quality and insomnia symptoms. In this subgroup, the prevalence of poor sleepers and clinical insomniacs decreased. Respondents preserving their screen time habits did not show any change in the sleep parameters.

**Conclusions:** Our investigation demonstrated a strong relationship between the modifications of the evening electronic device usage and the time course of sleep disturbances during the lockdown period. Interventions to raise public awareness about the risks of excessive exposure to backlit screens are necessary to prevent sleep disturbances and foster well-being during the home confinement due to COVID-19.

**Statement of Significance:** The present investigation is the first to provide insights about the relationship between the changes in evening electronic device usage and the time course of sleep disturbances during the COVID-19 lockdown. Consistent with the well-known negative effect of backlit screen lights on circadian physiology, we demonstrated a strong relationship between the screen time modifications in the hours before falling asleep, the development and exacerbation of sleep disturbances, and the changes of sleep/wake patterns during the period of home confinement due to COVID-19 pandemic. To date, hundreds of thousands of people are subjected to restraining measures worldwide. Our findings have large scale and broad-spectrum implications, considering the unavoidable increase of electronic device usage during the current period of limited social interactions.

## Introduction

The rapid worldwide spread of the COVID-19 pandemic marked the first months of 2020. During this unprecedented situation, governments across the globe implemented extraordinary measures to reduce the spread of the contagion and the pressure on the healthcare systems. From 9 March to 4 May 2020 a total lockdown was imposed in Italy, involving a large-scale closure of most work activities, social distancing, and a home-based quarantine imposition to the general population. The home confinement measures had a substantial negative impact on global mental health and psychological well-being.^1,2^ The considerable impairment of the daily routine had consistent repercussions on sleep health and circadian rhythms, as documented by several studies.^3–6^ However, evidence on the time course of sleep disturbances during the extended period of restraining measures is scarce.^7^ The forced social isolation and the limitations of the outdoor activities led to a worldwide increase in web-based social communication.^8–10^ Electronic devices daily usage increased^11–13^ to compensate for the limited social interactions, fill free time, and ward off boredom. The implementation of these habits may have helped to cope with the challenging and stressful isolation period. Nevertheless, the increase of screen exposure in the hours before bedtime could have determined adverse consequences on sleep health. The sleep rhythms are intimately linked with the ambient light, which represents a crucial regulator of the biological clock. Human eyes comprise non-visual photoreceptors that are primarily responsive to ∼450–480 nm light within the blue portion of the spectrum.^14,15^ The activation of this system leads to a suppression of the melatonin release, which is a key sleep-related pineal gland hormone.^16^ The evening exposure to short-wavelength-enriched light has alerting effects and detrimental consequences on sleep.^17,18^ Nowadays, most screens of modern electronic devices (computer, smartphone, tablet, television) are equipped with light-emitting diodes (LEDs) having a peak wavelength in the blue range of ∼460 nm.^19^ Therefore, the exposure to backlit screens during the hours preceding the habitual bedtime can interfere with the circadian physiology.

Epidemiological and cross-sectional studies indeed showed a strong relationship between the electronic device usage after the sundown and alterations of sleep patterns.^20–27^ The impact of screen exposure before falling asleep on the melatonin secretion was confirmed by several studies that experimentally manipulated the evening exposure to tablet,^28^ eReader,^29^ and computer screens.^19,30^ These investigations also reported decreased objective and self-reported sleepiness, higher sleep onset latency, and altered sleep architecture. Conversely, other studies showed protective effects of blocking blue light emissions on melatonin regulation^31^ and sleep quality, both in healthy^32^ and clinical insomniac subjects.^33,34^

Based on this evidence, the present study aimed to shed light on the relationship between the longitudinal changes of sleep disturbances between the third and the seventh week of home confinement in Italy and the retrospectively reported modifications of the exposure to electronic devices before falling asleep during the same lockdown period. We hypothesized that the changes in electronic device usage could be a crucial mediator of the lockdown-related sleep alterations over time. We expected that individuals that increased their screen exposure should have shown the largest sleep impairments and the most marked alterations of the sleep/wake schedule. On the other hand, subjects that reduced screen time should have exhibited a positive time course of sleep disturbances.

## Methods

### Participants and procedure

The present investigation is part of a larger research project aimed to understand the consequences of COVID-19 lockdown on the Italian population.^6^ A total of 7107 Italian citizens were recruited in a web-based survey through a snowball sampling during the third week of lockdown (25 March-31 March 2020). The survey comprised a demographic questionnaire (age, gender, education, occupation, and geographical location), the Pittsburgh Sleep Quality Index^35^ (PSQI), the Insomnia Severity Index^36^ (ISI), and the reduced version of the Morningness-Eveningness Questionnaire^37^ (MEQr), in this order. The PSQI is a validated tool to evaluate sleep quality comprising nineteen questions, from which a total score (range 0–19) is calculated. A score >5 identifies poor sleepers. The ISI is a validated instrument used to assess the severity of insomnia symptoms. The total score ranging between 0 and 21, and a score >14 indicates the presence of moderate/severe clinical insomnia condition. The MEQr is a validated questionnaire and its total score is used to identify the chronotype (4–10 score: evening-type; 11–18 score: intermediate-type; 19– 25 score: morning-type). Subsequently, respondents could decide whether continue the compilation of other three questionnaires (Beck Depression Inventory-second edition,^38^ BDI-II; 10-item Perceived Stress Scale,^39^ PSS-10; state-anxiety subscale of the State-Trait Anxiety Inventory,^40^ STAI-X1), with the option to stop after each of them. This optionality was aimed at ensuring higher reliability of the data collected, avoiding false answers in the last questionnaires. The BDI-II is a validated questionnaire used to assess clinical depression symptoms (range 0–63). The 10-PSS is a 10-item questionnaire evaluating the perceived stress following stressful events (range, 0–50). The STAI-X1 is a well-established 20-item scale measuring state anxiety (range, 1–80). For all these questionnaires, higher scores indicate more severe conditions.

After four weeks, the website link of the follow-up survey was provided to the participants via email address/telephone number. A total of 2701 subjects completed the second assessment in a seven-day period (21-27 April 2020). From this large follow-up sample, we included in the reported analyses only the 2123 respondents (mean age ± standard deviation, 33.1 ± 11.6; range, 18-82; 401 men, see Table 1) that completed the first survey during the four days preceding the daylight-saving time (25 March-28 March; Time 1). This allowed us to avoid interfering and confounding effects at the baseline measurement due to the summertime beginning (for a review,^41^). During the follow-up survey (Time 2), participants completed the same questionnaires of Time 1. Moreover, they were asked to retrospectively evaluate the changes (increase, maintenance, reduction) from the first assessment in the usage duration of electronic devices (smartphone, computer, tablet, television, eReader) in the two hours before falling asleep. The study has been approved by the institutional review board of the University of L’Aquila (protocol n. 43066) and has been carried out according to the principles established by the Declaration of Helsinki. Online informed consent to participate in the whole research was obtained from all the respondents during the first assessment.

**Table 1.** Sociodemographic composition of the sample participating in both the first and the second measurement (Time 1: 25-28 March 2020; Time 2: 21-27 April 2020).

|  |  |
| --- | --- |
| *Gender* |  |
| Male | 401 (18.9%) |
| Female | 1722 (81.1%) |
| *Age* |  |
| 18–30 years | 1263 (59.5%) |
| 31–50 years | 598 (28.2%) |
| > 50 years | 262 (12.3%) |
| *Education* |  |
| Until middle school | 31 (1.5%) |
| High school | 686 (32.3%) |
| Graduated | 1406 (66.2%) |
| *Occupation* |  |
| Unemployed | 184 (8.7%) |
| Employed | 1172 (55.2%) |
| Student | 767 (36.1%) |
| *Geographical location* |  |
| Northern Italy<sup>a</sup> | 767 (36.1%) |
| Central Italy<sup>b</sup> | 593 (27.9%) |
| Southern Italy<sup>c</sup> | 763 (35.9%) |
| *Electronic device usage* |  |
| Increased | 751 (35.4%) |
| Unchanged | 1221 (57.5%) |
| Reduced | 151 (7.1%) |

<sup>a</sup> Northern Italy: Aosta Valley, Emilia Romagna, Friuli-Venezia Giulia, Liguria, Lombardy, Piedmont, Trentino-Alto Adige, and Veneto. <sup>b</sup> Central Italy: Lazio, Marche, Tuscany, and Umbria. <sup>c</sup> Southern Italy: Abruzzo, Apulia, Basilicata, Calabria, Campania, Molise, Sardinia, and Sicily.

### Data analysis

To control for potential selection bias of the follow-up participants, we performed preliminary mixed model analyses comparing the Time 1 questionnaire scores of respondents that participated only to the first assessment and those who attended both the measurements (Time 1 and Time 2). These control analyses did not highlight significant differences (all P > 0.10).

According to the purpose of the present study, the main variables were the PSQI and ISI scores. Additionally, from the PSQI questionnaire, we extracted other variables such as total sleep time (TST, min), sleep onset latency (SOL, min), bedtime (BT, hh:mm), and rise time (RT, hh:mm). To evaluate the time course of the sleep dimensions as a function of the reported changes of exposure to electronic devices, all the above variables were submitted to mixed model analyses with a random intercept per participant, accounting for the expected intraindividual variability. The models comprised “time” (Time 1, Time 2), “screen exposure” (increased, unchanged, reduced), and their interaction as predictors. Additionally, “gender” (man, woman) was included as factor, and age as covariate, to control for putative effects of these demographic variables on the main outcomes of the present study. Subsequently, explorative analyses were carried out adding to the models the scores of MEQr, BDI-II, 10-PSS, and STAI-X1 as covariates. These further analyses aimed to control the effects of chronotype, depression, stress, and anxiety on sleep measures. Mixed model analyses were performed using the “lme4” R package.^42^ Models were fitted using REML, using the Satterthwaite approximation to compute P-values. Bonferroni post hoc tests were obtained using the “emmeans” R package.^43^ Finally, the validated cut-off scores of PSQI and ISI were used to determine the prevalence of poor sleepers and moderate/severe insomnia condition. Subsequently, McNemar tests were performed to evaluate the modifications of the prevalence of sleep disturbances between Time 1 and Time 2 in the three groups characterized by different changes of exposure to electronic devices before falling asleep. For all the analyses, statistical significance was set at P < 0.05, and all tests were 2-tailed.

## Results

### Relationships between screen exposure and sleep variables

Mixed model analyses did not highlight significant effects of the “time” factor for all the examined variables (PSQI: F_1,2027.32_ = 0.11, P = 0.75; ISI: F_1,2117_ = 0.75, P = 0.39; TST: F_1,2112.44_ = 1.37, P = 0.24; SOL: F_1,2112.92_ = 0.49, P = 0.49; BT: F_1,2111.55_ = 0.10, P = 0.76; RT: F_1,2111.77_ = 0.14, P = 0.71; respectively). “Screen exposure” was significant for all the variables (PSQI: F_2,2109.30_ = 29.57, P < 0.001; ISI: F_2,2116_ = 32.51, P < 0.001; TST: F_2,2115.08_ = 6.74, P = 0.001; SOL: F_2,2115.20_ = 6.14, P = 0.002; BT: F_2,2114.85_ = 7.14, P < 0.001; RT: F_2,2115.39_ = 5.26, P = 0.005). The interaction between “time” and “screen exposure” predictors (Figure 1 and Figure 2) was also significant for all the variables (PSQI: F_2,2010.42_ = 20.29, P < 0.001; ISI: F_2,2117_ = 23.70, P < 0.001; TST: F_2,2112.55_ = 9.07, P < 0.001; SOL: F_2,2113.04_ = 6.70, P = 0.001; BT: F_2,2111.64_ = 30.11, P < 0.001; RT: F_2,2111.84_ = 20.63, P < 0.001).

**Figure 1.**
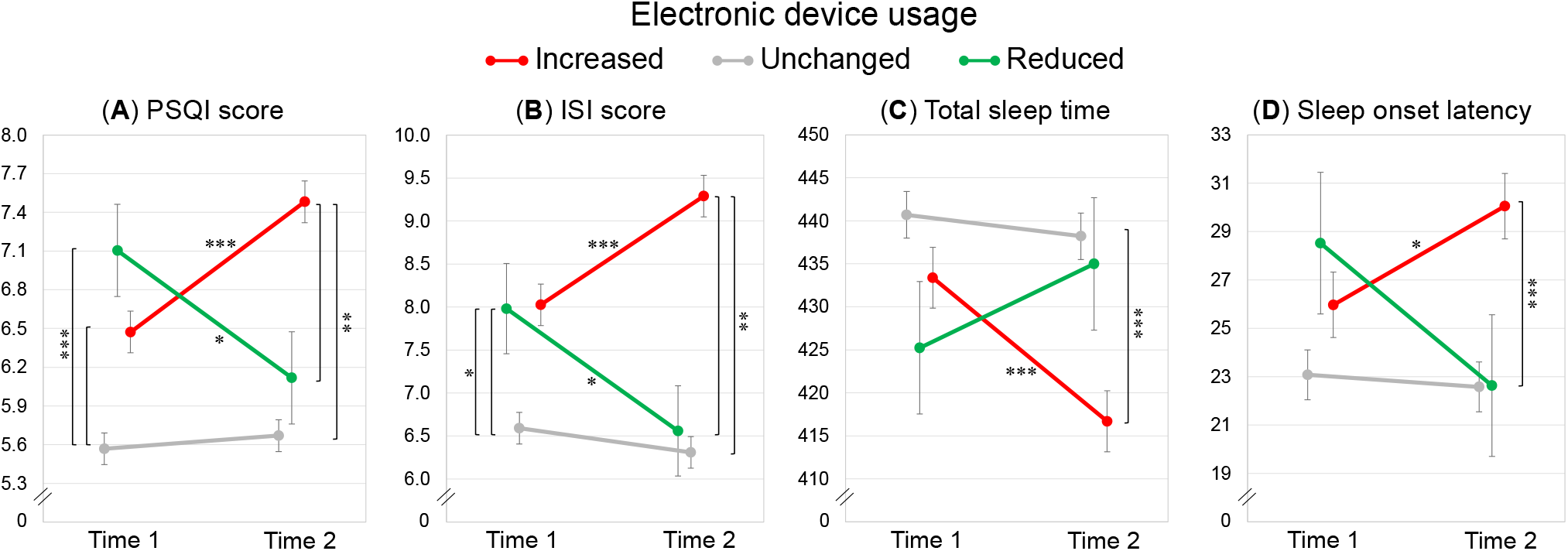
“Time” x “screen exposure” interaction for PSQI and ISI scores, total sleep time (min), and sleep onset latency (min). Mean ± standard error of the PSQI and ISI scores (A, B), total sleep time (C), and sleep onset latency (D) at the two assessments (Time 1, Time 2) for respondents who declared an increase, preservation, or reduction of the electronic device usage duration before falling asleep. Bonferroni significant post hoc comparisons are reported with asterisks (* P < 0.05, ** P < 0.01, *** P < 0.001).

**Figure 2.**
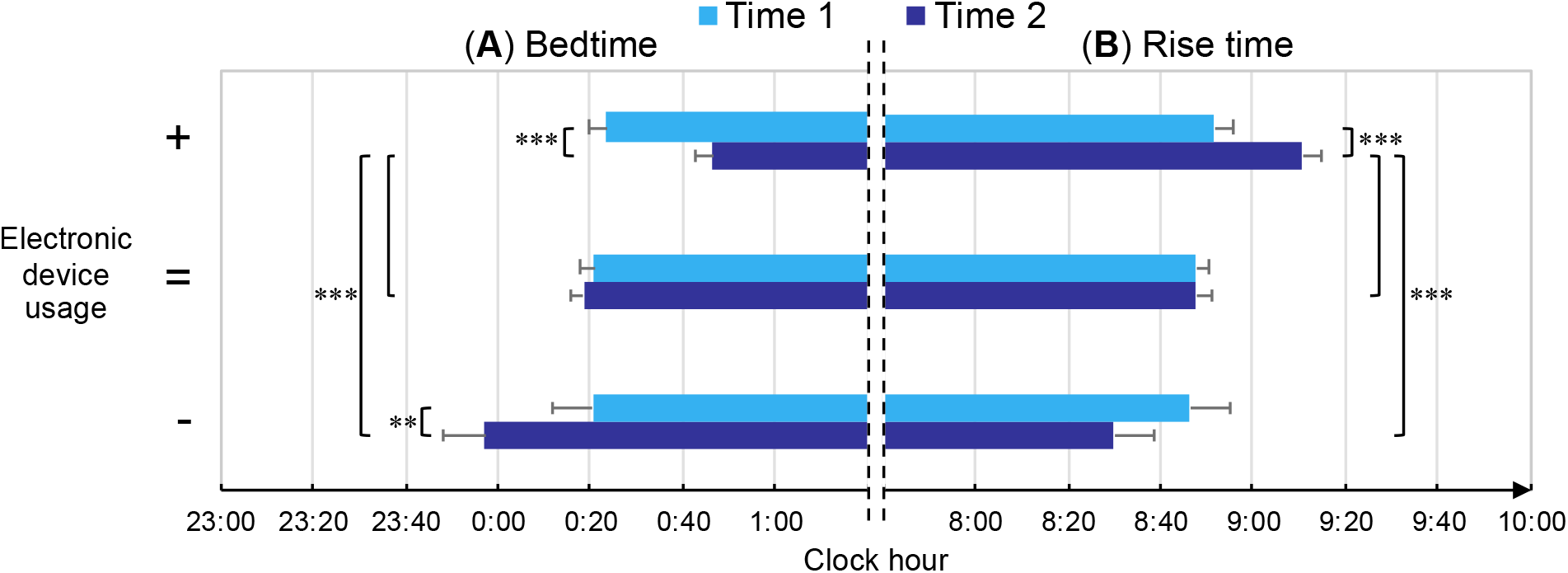
“Time” x “screen exposure” interaction for bedtime and rise time (hh:mm). Mean ± standard error of the bedtime (A) and rise time (B) at the two assessments (Time 1, Time 2) for participants who declared an increase (+), preservation (=), or reduction (-) of the electronic device usage duration before falling asleep. Bonferroni post hoc results are reported with asterisks (** P < 0.01, *** P < 0.001).

Post hoc comparisons between Time 1 and Time 2 suggested that participants who reported an increase of electronic device usage before falling asleep also showed a significant increase over time of PSQI and ISI scores (both P < 0.001), a reduction of TST (P < 0.001), a prolongation of SOL (P = 0.03), and delayed BT and RT (both P < 0.001). On the other hand, participants who reduced the screen exposure showed concurrent decreases of PSQI and ISI scores (P = 0.04, P = 0.02; respectively), an anticipation of the BT (P = 0.009), and a preservation of TST and SOL (P = 1.00, P = 0.58; respectively). No differences in all the variables were obtained for the participant who maintained unchanged their electronic device use habits (all P = 1.00).

At Time 1, there were no differences in PSQI and ISI scores between respondents who later reported an increase or a reduction of screen exposure (both P = 1.00). Participants maintaining their device use habits showed lower PSQI scores at Time 1, compared to those who increased or reduced the exposure to backlit screens (both P < 0.001). ISI scores were lower at Time 1 for subjects who did not change the screen exposure than participants who incremented or reduced it (P < 0.001, P = 0.04; respectively). The three groups did not differ at Time 1 on TST, SOL, BT, and RT (all P > 0.85).

Participants who reported an increase of screen exposure also showed higher PSQI and ISI scores at Time 2, and more advanced BT and RT compared to the other two groups (all P < 0.01), as well as shorter TST and longer SOL compared to the group that did not change the device usage habits (both P < 0.001, see Figure 2). No differences for all the variables were obtained at Time 2 between subjects who reduced or maintained the device usage duration before falling asleep (all P > 0.32).

Further control analyses confirmed the above-reported pattern of results, controlling for the covariance of age, gender, chronotype, depression, perceived stress, and anxiety. In particular, the interaction between “time” and “screen exposure” remained significant for all the variables (PSQI: F_2,1605.95_ = 17.50, P < 0.001; ISI: F_2,1685.08_ = 14.20, P < 0.001; TST: F_2,1694.81_ = 8.37, P < 0.001; SOL: F_2,1711.04_ = 4.53, P = 0.01; BT: F_2,1664.94_ = 17.11, P < 0.001; RT: F_2,1645.32_ = 12.70, P < 0.001), confirming the crucial mediation role of the changes in screen exposure in explaining the time course of the sleep outcomes during the lockdown. Finally, in light of the differences between the subgroups at Time 1 for PSQI and ISI scores, we performed control analyses to evaluate if the screen-related differences at Time 2 were affected by the baseline scores. ANCOVAs on the Time 2 ISI and PSQI scores with “screen exposure” as factor and Time 1 scores as covariate were significant (both P < 0.001). Bonferroni post hoc comparisons showed that respondents who increased the screen time showed the poorest sleep quality and the most severe insomnia symptoms (all P < 0.001) during the seventh week of lockdown, also controlling for the differences of the baseline scores.

### Relationships between screen exposure and sleep disturbance prevalence

McNemar tests highlighted a significant prevalence increase of poor sleepers (+11.4%) and of moderate/severe insomnia condition (+3.6%) in the group of respondents reporting an increased usage of electronic devices before falling asleep (χ^2^ = 108.23, P < 0.001; χ^2^ = 149.73, P = 0.01; respectively) (Figure 3). On the other hand, there was a significant decrease of poor sleepers (−10.7%) and of moderate/severe insomnia condition (−7.3%) in the group reporting a reduction of the device usage (χ^2^ = 19.90, P = 0.04; χ^2^ = 12.21, P = 0.04). Finally, in the group of participants who maintained their screen habits unchanged there was a reduction of clinical insomnia prevalence (−2.8%; χ^2^ = 188.51, P = 0.002), but not of poor sleepers’ prevalence (−2.3%; χ^2^ = 200.25, P = 0.17).

**Figure 3.**
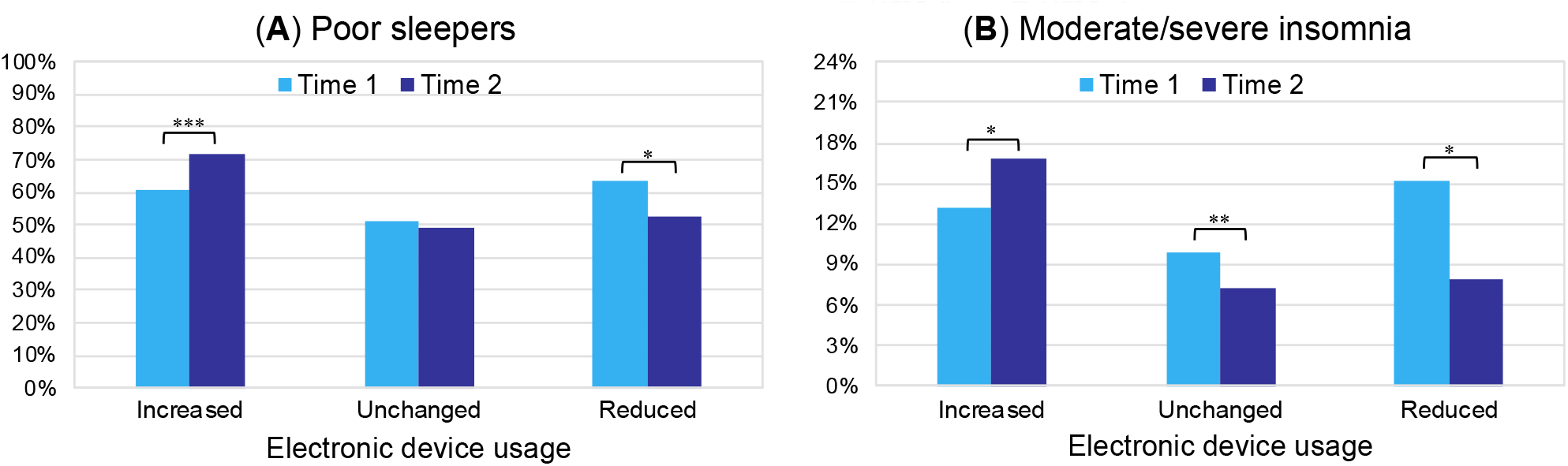
Prevalence of poor sleepers (A) and moderate/severe clinical insomnia condition (B) at the two assessments (Time 1, Time 2) for the respondents who increased, maintained unchanged, or reduced the usage of electronic devices before falling asleep. Significant results of the McNemar tests are reported with asterisks (* P < 0.05, ** P < 0.01, ** P < 0.001).

## Discussion

In the present study, we showed a strong relationship between the changes in evening screen exposure and the time course of sleep parameters during the COVID-19 lockdown. In line with the initial assumption, individuals declaring increased electronic device usage before falling asleep showed a general sleep impairment over time (from the third to the seventh week of home confinement). This outcome is exemplified by decreased sleep quality, an exacerbation of insomnia symptoms, reduced sleep duration, and longer sleep onset latency. Consistently, we found an increased prevalence of poor sleepers and moderate/severe insomnia condition only within this group of respondents. The increased screen exposure was also linked to delayed bedtime and rising time, outlining an advanced sleep phase across the home confinement period.

Furthermore, individuals who increased the device usage showed the poorest sleep quality, the most severe insomnia symptoms, the lowest sleep duration, the highest sleep onset latency, and they went to bed and woke up later compared to the other participants during the seventh week of lockdown.

On the other hand, participants reporting decreased evening screen exposure showed the opposite time course of sleep disturbances. They indeed exhibited improved sleep quality and mitigation of insomnia symptoms, which turned in a prevalence reduction of poor sleepers and clinical insomnia condition. This group of respondents anticipated bedtime after four weeks of home confinement. Finally, the respondents who maintained unchanged their electronic device usage habits did not show any modification in all the examined dimensions, except for a prevalence reduction of moderate/severe insomnia conditions.

Remarkably, we obtained the present findings controlling for the effects of gender and age, and they were confirmed also controlling for the covariance of chronotype, depression, stress, and anxiety. Therefore, our results indicate a direct relationship between evening device usage and time course of sleep disturbances during the home confinement period, independent of other psychological and circadian dimensions.

The pattern of results of our longitudinal investigation is consistent with a large pre-outbreak cross-sectional literature addressing the relationship between sleep and evening electronic device usage. In particular, higher screen time has been associated with reduced sleep duration,^20,44–46^ prolongation of sleep onset latency,^21–24,46^ later sleep onset and waking up,^20,21,45^ poor sleep quality,^20–24^ and insomnia symptoms.^21,26,44^

The COVID-19 pandemic has affected all the world, and the home confinement constitutes the most widely used measure to contrast the spread of the contagion. In modern societies, the increase of screen-based device usage could represent an unavoidable consequence of the pandemic-related home confinement periods. Indeed, more than one-third of our sample reported an increase in electronic device usage in the two hours before falling asleep. Consequently, our findings have substantial large-scale implications when contextualized to the current unprecedented situation.

Adequate sleep quantity/quality is essential to deal with stressful events^47^ and preserve mental health,^48,49^ and it plays a crucial role in emotional processing^50,51^ and mood regulation.^52^ Indeed, aberrant light exposure and excessive screen time were associated with sleep and mental health problems.53,54 Consistently, blocking screen-emitted blue light has proved to be effective in promoting at the same time sleep quality and mood^32,34^ and it was proposed as a useful approach to treat both clinical insomnia^33,34^ and mood disorders. ^53,55^ Finally, sleep and the circadian system support the proper functioning of the immune system,^56,57^ never as important as during the current historical period. In light of these considerations, the relationship between screen time and sleep outcomes has a broad spectrum of implications, configuring a major public health concern during COVID-19 outbreak.

The present results were obtained in an Italian sample, but they could be generalizable to other modern societies since the putative underlying mechanisms involve a disruption of circadian physiology and a direct arousal-induced effect caused by evening light exposure. However, we can not infer the causality of this relationship since this is an observational study, and the measurement of screen exposure changes has been retrospectively reported during the second assessment. Notwithstanding that comprehensive literature supported the detrimental effect of the blue light emitted by electronic devices on melatonin secretion and sleep patterns,^19,28–30^ we can not exclude reverse causation. Nevertheless, the two interpretations are not mutually exclusive, and a bidirectional model of causation has been suggested.^58^ We propose that a vicious circle during the confinement period was established, in which the increased screen exposure before falling asleep negatively impacted the sleep parameters, which in turn supported the overuse of electronic devices after the sunset.

In conclusion, our findings corroborate the assumption that the governments should pursue policies to raise public awareness on healthy sleep behaviors during the confinement due to the COVID-19 pandemic, discouraging the excessive use of electronic devices before falling asleep.^59,60^ The evening use of blue light glasses and the application of blue wavelength light filter (night shift settings) on the electronic screens should be encouraged to mitigate the well-known detrimental consequences of electronic device usage. To date, the feared risk of a second wave of contagion is increasingly becoming a concrete reality, and hundreds of thousands of people are subjected to home confinement measures worldwide. In light of our results, the above-mentioned interventions focused on sleep hygiene are fundamental to counteract the occurrence and exacerbation of sleep disturbances and foster the general well-being during the home confinement due to the COVID-19 pandemic.

### Limitations

To the best of our knowledge, the present investigation is the first to provide insights about the relationship between electronic device usage and the time course of sleep disturbances during the COVID-19 lockdown. However, it should be acknowledged that we used a non-probabilistic sampling technique, and the sample comprised a higher prevalence of women and young people. Moreover, under-eighteen years-old individuals were not included. However, the relationship between evening screen time and sleep disturbances was widely shown in adolescents.^61–63^ We hypothesize that our results could be generalizable to the youngest people. Additionally, the electronic device category of our survey included a broad set of devices, and we can not discern the relationship between each device usage and the time course of the sleep outcomes. Finally, in our survey, we did not assess the implementation of protective approaches to reduce the emission or perception of screen light, and thus we can not estimate their contribution to the present findings.

## Data Availability

The data underlying this article will be shared on reasonable request to the corresponding author.

## Acknowledgments

We are grateful to Professor Marco Lauriola for his valuable support in the statistical analysis and to Jasmin Cascioli for her help in data collection.

## Author contributions

Conceptualization, F.S. and M.F.; Methodology, F.S. and M.F.; Investigation, F.S., G.A., D.C., L.V.; Data curation, F.S.; Formal analysis, F.S.; Writing – original draft, F.S.; Writing – review & editing, F.S., A.D.A., D.T. and M.F.; Supervision, M.F.

